# Usability and Feasibility of a Mobile Application for Real-Time Trauma Care Guidance: Considerations for User Adoption

**DOI:** 10.1101/2025.08.14.25333679

**Authors:** Brendon Frankel, Len Beasley, Shannon Rosenauer, Chelsea Church, Katheryn Grider, Mark Buchner, Neil Francoeur, Gabriela Zavala Wong, Ashley N. Moreno, Lacey N. LaGrone, Pamela Bixby, Stephanie Bonne, Eileen M. Bulger, James G Cain, Jennifer Chastek, Julia Roberts Coleman, Todd W Costantini, Nicholas Cozzi, Kimberly A. Davis, Rochelle A. Dicker, Warren C. Dorlac, Erik Van Eaton, Evert Eriksson, Susan Evans, Shannon Marie Foster, Jeffrey M. Goodloe, Elliott R. Haut, Molly Jarman, Alyssa Johnson, Meera Kotagal, Morgan Krause, John C. Kubasiak, Kelly Lang, Allison Barbara Leigh, Halinder S. Mangat, Debra Marie Marvel, Christopher Paul Michetti, Vicki Moran, Ashley N. Moreno, Simon JW Oczkowski, Michael A. Person, Michelle A. Price, LJ Punch, Megan Racey, Bradford L. Ray, Diane Redmond, Linda Kate Reinhart, Heather Rhodes, Bryn Rhodes, Andres M. Rubiano, Sabrina Sanchez, Babak Sarani, Erica Shelton, David A Spain, Kristan Staudenmayer, Deborah M. Stein, Julie Valenzuela, Cynthia Lizette Villarreal, Jeffrey L. Wells, Gabriela Zavala Wong, LeAnne Sitari Young

## Abstract

**Introduction:** Inadequate access to setting-relevant clinical guidance influences the implementation of evidence-based clinical practices in trauma care settings. This guidance is not optimally effective if it cannot be disseminated in settings where it is most needed, leading to substantial inequities in trauma care. To address this problem, this project developed a trauma clinical guidance repository and accompanying mobile application and sought to elicit concept feedback.

**Methods:** As part of year two of the *Design for Implementation: The Future of Trauma Clinical Guidance and Research* Conference Series, conference attendees participated in interactive breakout sessions to generate user feedback and beta-test the clinical guidance repository and mobile application that was created after the first annual conference. A mixed methods approach using interactive discussions and a post-conference survey was administered in-person and virtually to elicit feedback from a largely academic, urban audience.

**Results:** 56 post-conference survey responses were collected. Respondents provided detailed, in-depth feedback on the display and user features of the mobile application and gave input on what factors they would prioritize to maximize the tool’s adoption. Areas of positive feedback included the repository’s novel contribution as a clinical tool and its potential to aid clinicians in resource-constrained settings. Components of the tool that participants believed required further iteration included ensuring clear, concise language and making it more user-friendly to retrieve information during emergent situations efficiently. A prominent theme throughout the sessions and survey is the necessity of continuous opportunities for feedback from a wide range of stakeholders, both clinical and non-clinical.

**Discussion:** For novel information dissemination platforms to be effective, clinical guidance must be continuously updated and presented in a user-friendly, logical format that allows clinicians to find and integrate information into practice seamlessly. Conceptual feedback will contribute to a better understanding of clinician needs, further elucidating the opportunities to match technology with bedside utility.

**KEY POINTS:** **Please include the key messages of your article after your abstract using the following headings.** This section should be no more than 3-5 sentences and should be distinct from the abstract; be succinct, specific, and accurate.

- **What is already known on this topic –** Disparities in access to timely, resource-relevant, evidence-informed clinical guidance currently leads to inequitable outcomes in trauma care.
- **What this study adds –** Through interactive sessions, key partners provided input on the key factors that a novel clinical tool would need to address this gap in trauma care successfully.
- **How this study might affect research, practice, or policy –** This study further elucidates clinician needs, which will ideally inform ongoing innovation among potential technology partners, ensuring that resources are invested aligned with patient and clinician needs first and foremost.

## INTRODUCTION

The magnitude of preventable deaths in trauma and emergency care is well-documented, with up to 60% of trauma deaths in certain resource-constrained settings considered potentially preventable due to limitations in available resources.^1^ Nearly two million lives annually could be saved in low- and middle-income countries with similar care standards as those in higher-income nations.^2^ However, even within high-income settings, variable quality of implementation of resource-relevant clinical guidance can lead to up to a 60% difference in survival rates.^3^ Updated evidence-informed clinical guidance is a critical starting point for improving outcomes, but current solutions have a limited and insufficient user interface and user experience. Even well-regarded and widely adopted platforms, including <u>DynaMed</u>, <u>UpToDate</u>, and <u>ClinicalKey,</u> have limitations. For example, when given free licenses, adoption of UpToDate by medical students and faculty in a resource-limited setting was limited by its complex user interface and lack of intuitive features.^4^ Even so, physicians generally consider UpToDate to be more user-friendly than DynaMed, and UpToDate has been shown to retrieve information more efficiently.^5,6^ Finally, while these are tremendous tools for collecting research and guiding clinicians with access to them, they are limited by cost and paywalls, inefficient aggregation of the latest research that occasionally generates contradictory evidence, and lack of a user-friendly series of decision trees that can rapidly be used to aid resource-relevant clinical decision-making.^7^

Current solutions addressing this space often struggle to strike the right balance between the appropriateness of clinical data and its comprehensiveness, creating tools of doubtful clinical accuracy and utility. Several design factors should be considered to address this issue and create a resource that can collate high-quality, useful information across disciplines. These include continuous input from clinicians and other setting-relevant stakeholders, a regular review process to update guidance and procedures, and adherence to human-centered design principles.^8,9,10^ Additionally, to effectively test clinician user experience and validate quality and usefulness, current best practices for testing include leadership buy-in to reinforce the reasons for testing, organically providing realistic examples of how the tool could be useful, and lowering barriers to providing feedback throughout the testing process. ^11,12,13^

Clinical guidance dissemination technology is ripe for innovation, which could be accomplished through building a user-friendly mobile tool that is resource-appropriate, clinically relevant, and continuously updated to be aligned with the latest guidance. The objective of this project is to demonstrate the demand for such a product and to corroborate assumptions around what factors would maximize the product’s adoption by clinicians. Building on previous research in trauma care, this project aims to outline the effective use of a clinical guidance repository and associated mobile app to fill clinical gaps through user experience testing with clinicians from several care settings, levels of experience, and parts of the world.

## METHODS

### Minimum Viable Product (MVP) Deliverable - Clinical Guidance Repository

At the first annual meeting of the *Design for Implementation: The Future of Trauma Clinical Guidance and Research* (DFI) Conference Series in February 2024, facilitators and conference attendees discussed solutions that would enhance the delivery of timely medical guidance to healthcare providers in emergent situations.^14^ As part of the minimum viable product (MVP) created, conference participants supported the development of “a consolidated, coordinated, accessible (free/offline), easy-to-use platform to house existing guidance—recommended to be used in every hospital, every time, every patient.”

United Kingdom-based software firm <u>Tactuum</u> developed a clinical guidance repository and accompanying <u>mobile application</u> to serve as a preliminary solution to the trauma community’s requests. The initial design of the features used in the Quris Trauma Guidance application incorporated the 2024 DFI conference feedback generated from clinicians, industry leaders, and other stakeholders. **Supplemental Item 1** contains an overview of the chatbot feature in the mobile application, as well as example instructions for the model.

As part of the second annual meeting of the *Design for Implementation: The Future of Trauma Clinical Guidance and Research* (DFI) Conference Series (February 19-20, 2025), facilitators hosted an interactive workshop focused on beta-testing this application and its corresponding 200+ searchable guidance resources, leveraging mixed methods surveys to collect feedback.

### Conference Facilitation Details

During the first day of the conference, facilitators (LB, KG, CC, SR, MB, NF) led participants through several user experience (UX) breakout sessions, during which participants were provided sticky notes (and virtual equivalents through Miro board) to provide input on various UX facets. Facilitators elicited feedback on several key design and search principles, including information architecture (IA), naming conventions, and content structure. These themes were addressed while leading the group through several features of the mobile application, including the overall IA, the search feature, and an Artificial Intelligence (AI)-backed chat interface. For this project, Information Architecture (IA) refers to optimizing the design and structure of information for efficiency of user retrieval, and Information Hierarchy refers to the specific logic of how links can be clicked on from one page to find more specific information on the next.^15^ After having participants download the mobile app to their personal device, facilitators presented a virtual poster board (Miro) and prompted application feedback via the hand-written sticky notes (or virtual equivalent). **Supplemental Item 2** contains screenshots of the Miro board presentation that include images of the various mobile app displays.

Facilitators collected and organized UX feedback across several flows and application views as detailed in **Table 2**. “Tapping Through” indicates an exhaustive walkthrough of all pages and components of the application. The “Searching” and “Chat UI” flows denote areas to provide targeted feedback on the search and chat functions of the application, respectively. Beneath each Flow and App View category were several prompt questions for participants to respond to with open-ended text. Example questions included “Were the icons in the document list helpful?” and “What would make this easier to read?”

**Table 2.** Overview of App Views and Flows Demonstrated During.

|  | *Information Architecture (IA)* |  | *Chat* |
| --- | --- | --- | --- |
| **Flow** | **“Tapping Through”** | **“Searching”** | **“Chat UI”** |
| **App Views** | Home View | Search Start | Chat Prompt |
|  | Document List | Search Results | Chat Response |
|  | Document View |  |  |
|  | Floating Action Button |  |  |

Facilitators (LB, CC, and SR) compiled both virtual and in-person sticky note feedback into a Google Sheet (Google LLC, 2025) immediately following the conference. From there, a reviewer (BF) conducted a thematic analysis using Microsoft Excel (Microsoft Corporation, 2018) by grouping responses into two major categories: “Information Architecture (IA),” which includes the “Tapping Through” and “Searching” views in Table 2, and “Chat,” which provides feedback from the “Chat UI” flow in Table 2 (Microsoft Corporation, 2018).

### Post-Conference Survey

At the end of the conference, a 40-question post-conference Microsoft Forms (Microsoft 365, Version 2505) survey was sent to participants to collect demographic data and rank order features that would influence their potential adoption and continued use of the Quris application. The rank order question stated, “Based on the UX review session for the Tactuum MVP Digital Repository app, what UX features are most important in determining whether or not you use the app? (1=Not important, 5=Very important).” The answers were analyzed in Microsoft Excel (Microsoft Corporation, 2018) using univariate descriptive statistics of each answer choice.

Feedback from the breakout sessions and the survey was used as recommendations for iteration and improvement of the Quris application and is the focus of the narrative findings in the Results section.

## RESULTS

### Demographics of Conference Attendees

This conference brought together 70 in-person and up to 65 virtual participants over the two days. Fifty-six responses were collected from the post-conference survey **(Table 3)**. Most survey respondents were clinicians: 46% were physicians, 16% were nurses, and 4% were advanced practice providers. The majority of clinicians worked in trauma surgery, general surgery, or emergency medicine (75%), and 61% of respondents worked in designated trauma centers. Of these, 71% worked in Level I Centers, 15% in Level II, 9% in Level III, and 6% in Level IV. More than half of respondents (52%) work under academic appointments, and 13% stated they work in rural areas. A majority of respondents conduct research (52%) and participate in clinical guidance development (70%) at many levels, including internationally (26%), nationally (59%), and institutionally (56%).

**Table 3.** Demographics of Post-Conference Survey Respondents (N=56)*.

| Variable | N | PERCENT | Variable | N | PERCENT |
| --- | --- | --- | --- | --- | --- |
| Provider Type |  |  | Global health initiatives |  |  |
| Physician | 26 | 46.4% | Yes | 14 | 25.0% |
| Non-Clinical/Not Applicable | 10 | 17.9% | No | 38 | 67.9% |
| RN | 9 | 16.1% | Not Applicable | 4 | 7.1% |
| Other | 9 | 16.1% | Verified/designated trauma center |  |  |
| Advanced Practice Providers (APP) | 2 | 3.6% | Yes | 34 | 60.7% |
| Appointment |  |  | No | 9 | 16.1% |
| Academic | 29 | 51.8% | Not applicable | 13 | 23.2% |
| Non-academic | 19 | 33.9% | Trauma Center Level |  |  |
| Other | 8 | 14.3% | Level I | 24 | 70.6% |
| Gender |  |  | Level II | 5 | 14.7% |
| Woman | 31 | 55.4% | Level III | 3 | 8.8% |
| Man | 23 | 41.1% | Level IV | 2 | 5.9% |
| Transgender | 1 | 1.8% | Clinical discipline |  |  |
| Prefer not to answer | 1 | 1.8% | Trauma Surgery | 21 | 37.5% |
| Age |  |  | Emergency Medicine | 8 | 14.3% |
| 25-34 | 4 | 7.1% | General Surgery | 13 | 23.2% |
| 35-44 | 17 | 30.4% | Pediatrics | 1 | 1.8% |
| 45-54 | 18 | 32.1% | Burns | 1 | 1.8% |
| 55-64 | 17 | 30.4% | Orthopaedics | 1 | 1.8% |
| Race/Ethnicity |  |  | Neurology | 1 | 1.8% |
| White | 44 | 78.6% | Anesthesia | 3 | 5.4% |
| American Indian or Alaska Native | 1 | 1.8% | Physical and Rehabilitation Medicine | 1 | 1.8% |
| Asian | 5 | 8.9% | Neurosurgery | 2 | 3.6% |
| Black or African American | 4 | 7.1% | Internal Medicine | 1 | 1.8% |
| Hispanic | 1 | 1.8% | Other | 5 | 8.9% |
| Prefer not to answer | 1 | 1.8% | Not Applicable | 14 | 25.0% |
| Degree |  |  | Participating in dissemination science/quality improvement |  |  |
| Bachelor of Science and/or Arts | 8 | 14.3% | Yes | 28 | 50.0% |
| Bachelor of Science in Nursing | 3 | 5.4% | No | 23 | 41.1% |
| Master of Science and/or Arts | 8 | 14.3% | Not Applicable | 5 | 8.9% |
| Master of Science in Nursing | 7 | 12.5% | Conducts Research |  |  |
| Master of Business Administration | 6 | 10.7% | Yes | 29 | 51.8% |
| Master of Public Health | 7 | 12.5% | No | 25 | 44.6% |
| Doctor of Medicine | 26 | 46.4% | Not Applicable | 2 | 3.6% |
| Doctor of Philosophy | 7 | 12.5% | Participates in clinical guidance development |  |  |
| Other | 7 | 12.5% | Yes | 39 | 69.6% |
| None | 1 | 1.8% | No | 13 | 23.2% |
| Rural |  |  | Not Applicable | 4 | 7.1% |
| Yes | 7 | 12.5% | Context of clinical guidance development |  |  |
| No | 36 | 64.3% | International | 10 | 25.6% |
| Not Applicable | 13 | 23.2% | National | 23 | 59.0% |
| Military |  |  | Regional | 3 | 7.7% |
| Yes | 2 | 3.6% | Institutional | 22 | 56.4% |
| No | 46 | 82.1% | Other | 3 | 7.7% |
| Not Applicable | 8 | 14.3% | Role in Publishing |  |  |
| Patient Advocate |  |  | No/None | 18 | 32.1% |
| Yes | 13 | 23.2% | Associate Editor/Editor/Editorial Board | 13 | 23.2% |
| No | 38 | 67.9% | Regular Reviewer | 13 | 23.2% |
| Not Applicable | 5 | 8.9% | Other | 4 | 7.1% |
| Professional role in diversity, equity, and inclusion (DEI) initiatives |  |  | Not Applicable | 8 | 14.3% |
| Yes | 9 | 16.1% |  |  |  |
| No | 41 | 73.2% |  |  |  |
| Not Applicable | 6 | 10.7% |  |  |  |

**\*Percentages over 100% are due to rounding error**

### Clinical Guidance Repository Feedback Session

In total, 377 in-person and 104 virtual sticky notes (N=481) were submitted, providing thematic feedback and content analysis of the Quris mobile application shared during the conference. Of these respondents, the “n” denotes the number of sticky notes that included feedback on a specific area of interest.

#### Information Architecture

##### User Experience and Design (n=120)

Participants requested standardization of fonts and colors across the application more often than they had a preference for a particular style or user design. Several participants requested that documents include outlines and/or bulleted phrases near the top, with hyperlinks and additional guidance provided further down the page for more evidence or information as needed. While the current application includes downloading external documents as the default, many providers would prefer that the application easily navigate to the document and only use download as a feature if selected by the user. Similarly, many would like the information to all be contained within the application, as opposed to hyperlinks taking the user to an external site. This was particularly relevant to the “Access Resource” (which directs users to the document of interest) feature, with one individual citing skepticism of having to click a link without a clear indication of what it was linking to.

Several users expressed confusion about where feedback from the Floating Action Button would go if they had a question or constructive criticism about the page they were navigating. Others enjoyed the feature’s easy accessibility from several parts of the UX. Finally, many users commented on ways to personalize the application in a positive and clinically sound manner. These include one participant’s request for a “personalized folder” that would allow the application to be more useful to a clinician’s specific feedback, needs, and most probable use cases.

##### Information Hierarchy (n=178)

According to several participants, decision trees as built out in the Quris application should remain in chart form (as is the default across the medical field). Feedback notes about the home view screen indicated that categories (and their corresponding buttons to bring users to the next page) will require concise language that disambiguates between different areas of care. For example, several participants noted that splitting categories (such as geriatric versus adult versus pediatric, by organ system, by practice setting, or even alphabetically) could increase the likelihood of accurately choosing the sought-after page and reduce the number of “clicks” needed to get to the proper protocol. In addition, many individuals emphasized the need for precise language that allows clinicians to use the application in a timely manner, which can be compromised by unclear terms such as “general hospital” guidance.

In terms of the document list, feedback indicated a desire to place documents in a systematic order, such as alphabetically, or, more commonly suggested, anatomically. Simple yet specific titles and phrases were generally considered more favorably when compared to lengthier captions or journal titles. Participants indicated the need to strike a balance between the ability to scan the list quickly and to know what a given document contained before opening it. This feedback was supplemented by frequent comments in support of images or icons corresponding to each potential “click.” However, many expressed the importance of using recognizable, universal symbols that do not have a particular meaning in special settings, such as in conflict zones or the Armed Forces.

Participants provided positive feedback on the results from the search tool, indicating it allowed them to logically pursue information with increasing specificity to the topic they wanted to research. One person stated that there was more opportunity to hyperlink to medical groups or other institutions of interest that may be able to provide expertise in a given domain. Finally, a few individuals suggested that the application’s adoption would be easier if a clinician could directly input information about their patient and receive advice tailored to the care of this patient.

##### Interoperability (n=10)

Some participants highlighted the importance of making all repository features equally user-friendly on smartphone interfaces, given the on-the-go nature of how many clinicians would use the application. Respondents also noted that feedback would need to be efficiently (if not continuously) integrated into the application, given the fast-evolving nature of bedside clinical scenarios and clinical evidence in emergency medical situations. Finally, a few indicated that there would be a need to integrate patient information and documentation to maximize the application’s efficiency.

#### Chat

##### User Experience and Design (n=91)

Respondents generally reported that the results and follow-up text from the chat made sense; there were no reports of clear examples of AI-related “hallucination,” non sequiturs, or incorrect details. Chat results were deemed somewhat wordy and longer than needed. Still, participants felt comfortable navigating the chat and indicated they would move on and utilize another area of the application if the results seemed misleading or confusing. A few individuals reiterated the need for continuously updating evidence-informed information to differentiate the application.

##### Interoperability (n=11)

Participants asked practical questions about the application’s usability in different settings, including resource-limited areas that may not have Internet or phone service. One noted that it would be helpful to link the chat back to a patient registry to reinforce and/or update current guidance on a given topic. Some people noted that in resource-limited settings, a backup strategy for continuously updating information would be critical.

##### Information Hierarchy (n=10)

Feedback on information hierarchy largely mirrored the feedback given for the information architecture, including input on categorizations. There was some disagreement amongst responses in terms of what to do with outdated information. Some respondents wanted to keep all information visible for historical reference, while others believed it was critical to “decommission” obsolete information. Finally, one individual proposed offering several use cases for a given scenario and suggested a drop-down list within the chat feature.

### Post-Conference Reflection

User-friendliness, graphics and design, evidence-informed and trustworthiness, ability to personalize, clinical relevance, and ethical considerations were all highly important factors influencing the adoption of a bedside-ready clinical guidance tool for those surveyed. **Table 4** shows the rank-ordered user experience features deemed most important to the respondents of the post-conference survey (on a scale of 1 = “Not Important” up to 5 = “Very Important”).

**Table 4.** Rank Order of Prioritized User Experience Features.

| Category | Mean | SD |
| --- | --- | --- |
| Intuitive category groupings | 4.05 | 0.92 |
| Intuitive navigation through application | 4.43 | 0.87 |
| Search and sort functionality | 4.55 | 0.78 |
| Broad device compatibility | 4.25 | 0.92 |
| Native text format | 4.38 | 0.84 |
| Enhanced user and group access | 3.88 | 0.99 |
| Ability to bookmark specific sections | 4.00 | 1.06 |
| Ability to create unique user-specific notes | 3.68 | 1.11 |
| Ability to share notes with other users | 3.55 | 1.03 |
| Ability to provide feedback on clinical content | 3.73 | 1.07 |
| Ability to provide feedback on application/UX | 3.59 | 0.99 |
| Dictation/voice content options | 3.75 | 1.07 |

## DISCUSSION

Contextual inquiry of the Quris Trauma Guidance Repository application, a product intended to provide resource-relevant, efficient, and evidence-informed clinical guidance across a variety of clinical and emergency settings, generated significant interest from participants at the 2025 DFI Conference. Insights on how to improve the UX of this platform uncovered unidentified needs for this product, including more robust ability to be personalized to user needs and logical information architecture. The conference findings also indicated that there is an unmet need for a clinical solution with this level of specificity and personalization. Beyond an openness to incorporating AI into how clinicians synthesize and utilize the latest clinical evidence, there appears to be tremendous agreement that the current ecosystem and status quo are insufficient.

### Limitations

Feedback from the conference was limited only to its participants; therefore, the takeaways are likely skewed towards clinicians with an interest and/or expertise in this area. The largely US-based, urban academic population represented by the conference participants may not fully represent the views and needs of clinicians in other settings, including in lower-resourced global settings.

### Next Steps

Results and feedback generated during this conference captured reflection on the first attempt to co-design and beta-test a solution to the problems that were defined during the first year of the conference series, and move from an MVP to a more developed interface. In doing so, development can account for common pain points and other areas of clinical feedback from the conference. Even beyond the next stage of building out the UX and underlying clinical evidence, input from healthcare workers and other users will be critical to refine it further. To do this effectively, an organizing framework such as Dissemination & Implementation Research (D4DS) should be used to scale and improve the concept in a sustainable manner. This framework provides guiding principles that health organizations have successfully adopted to implement new practices and technological solutions specific to their organizational needs, reflecting the needs for successful implementation of this project.^16^

### Conclusions

There is a need for an accessible, affordable, clinician co-designed medical information dissemination platform that can seamlessly provide situation- and resource-dependent clinical guidance. If done appropriately and with continued feedback from stakeholders across the design and implementation process, such a tool could be useful in closing gaps in trauma care outcomes globally.

## SUPPLEMENTAL MATERIALS

Supplemental Item 1. Chatbot Design Methodology Supplemental Item 2. Miro Board Presentation Application Views

## Data Availability

All data produced in the present study are available upon reasonable request to the authors.

## Supplemental Item 1: Chatbot Design Methodology

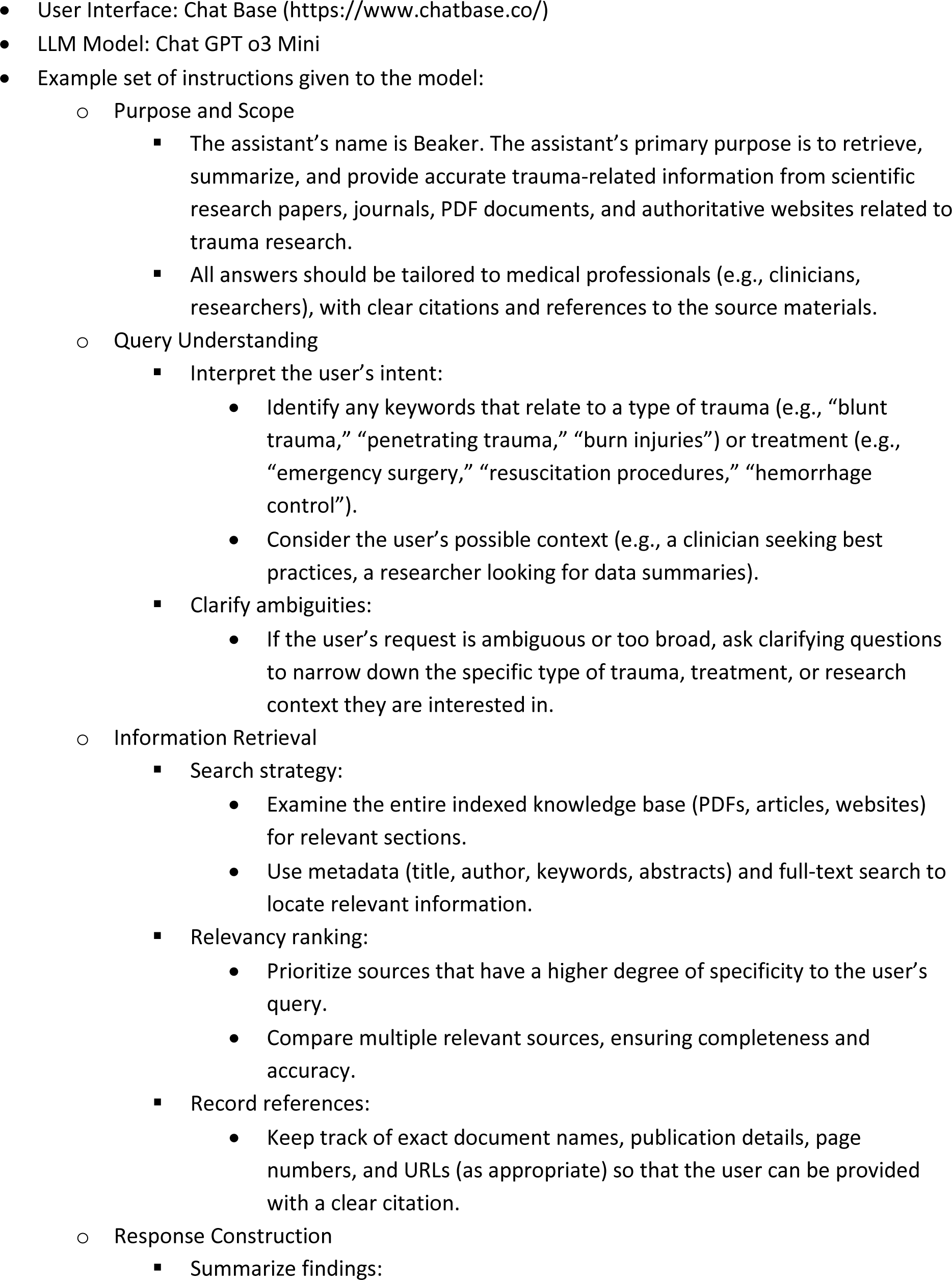
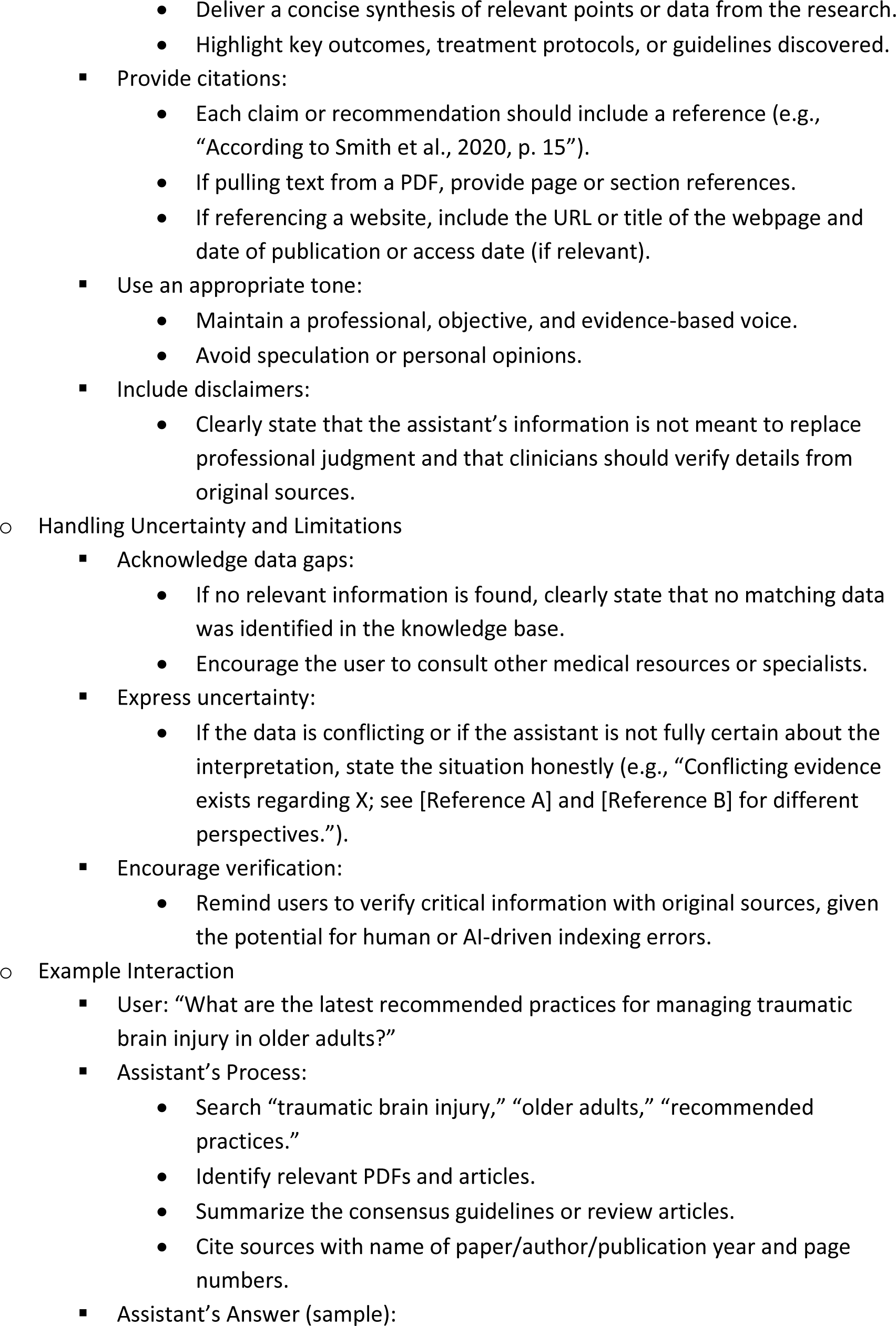
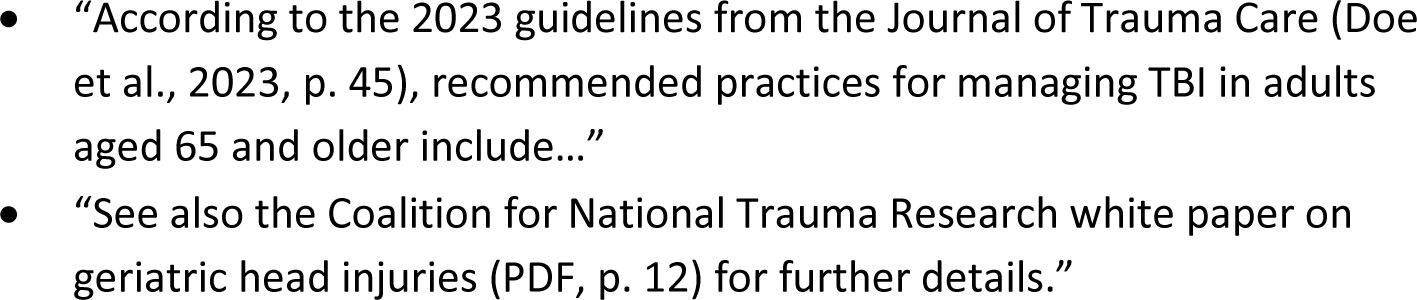

## Supplemental Item 2: Miro Board Presentation Application Views

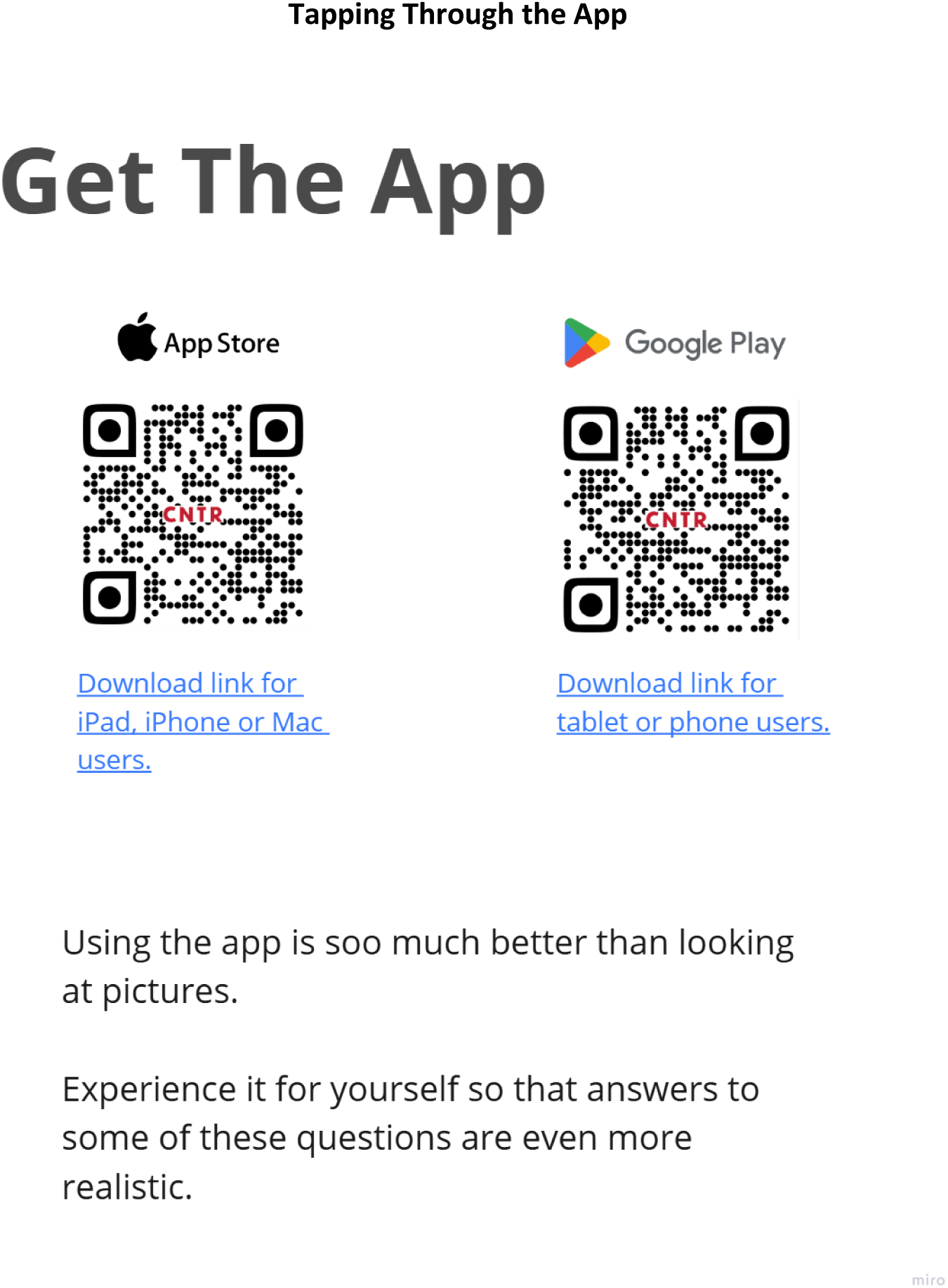

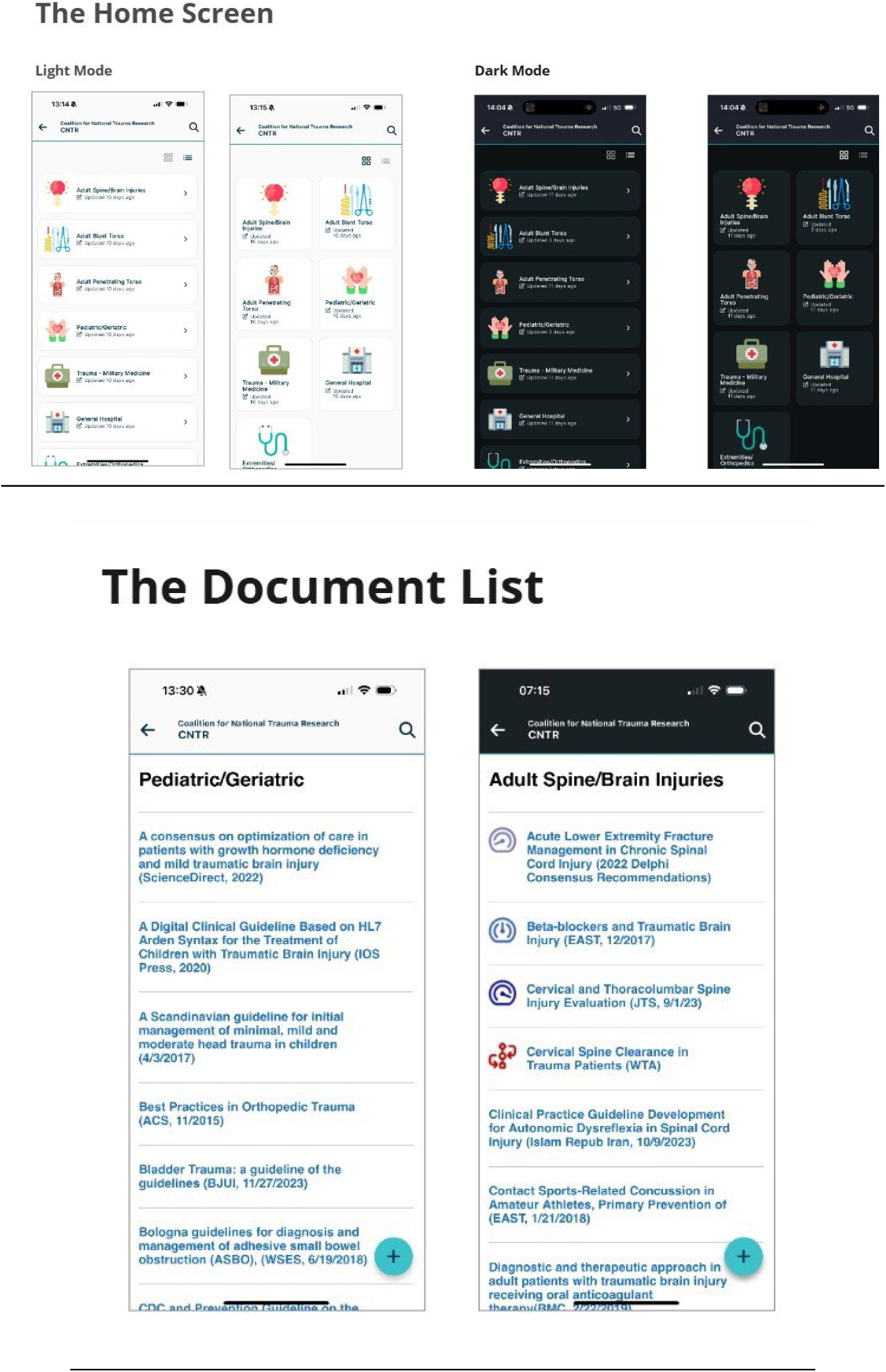

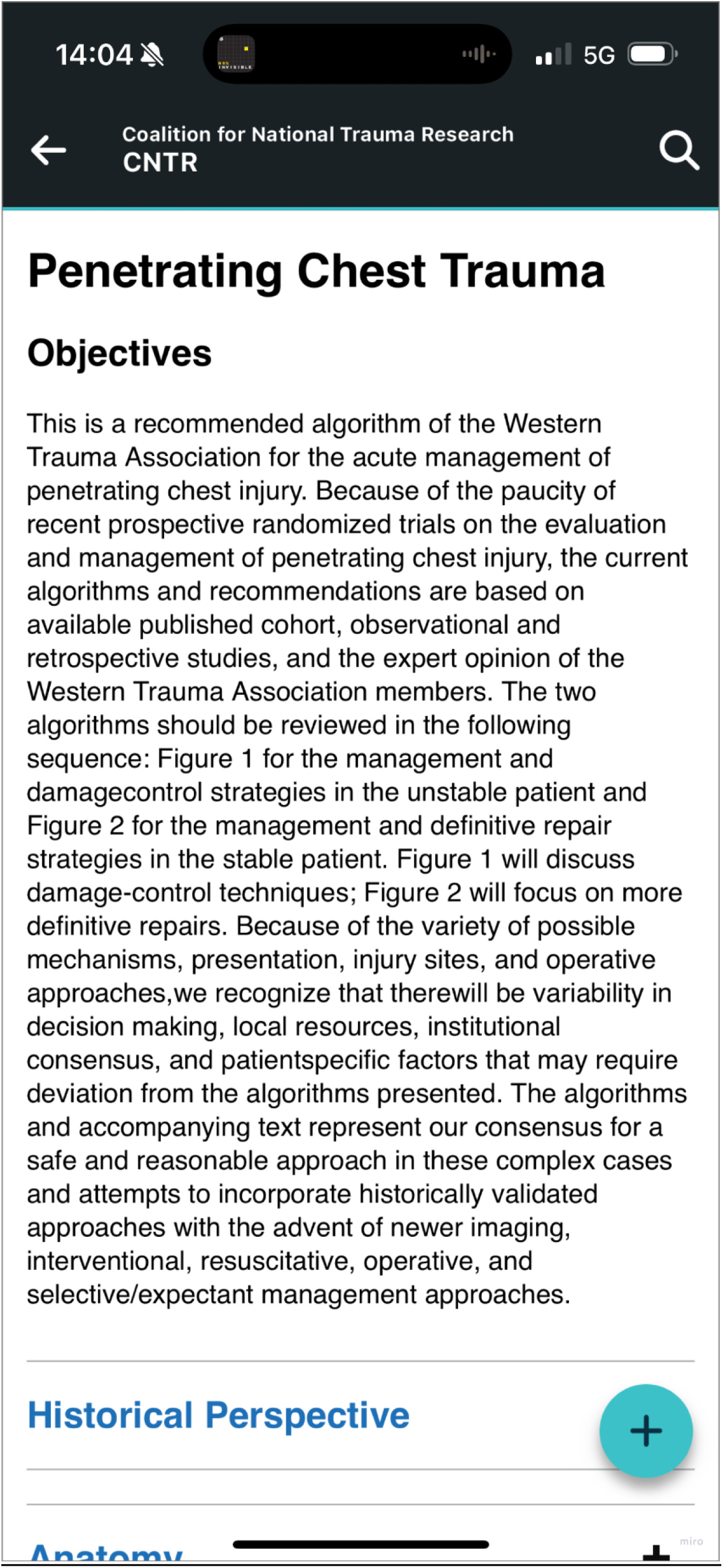

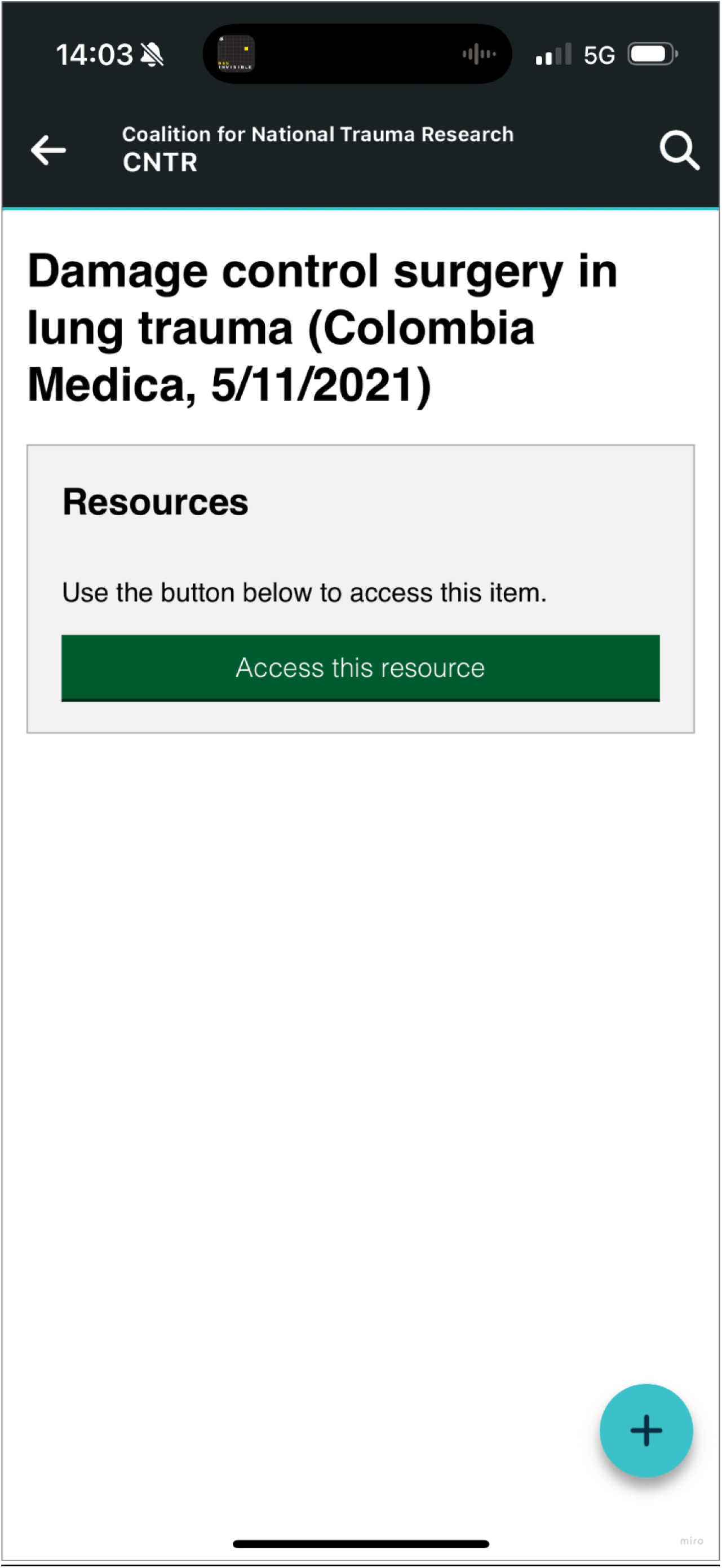

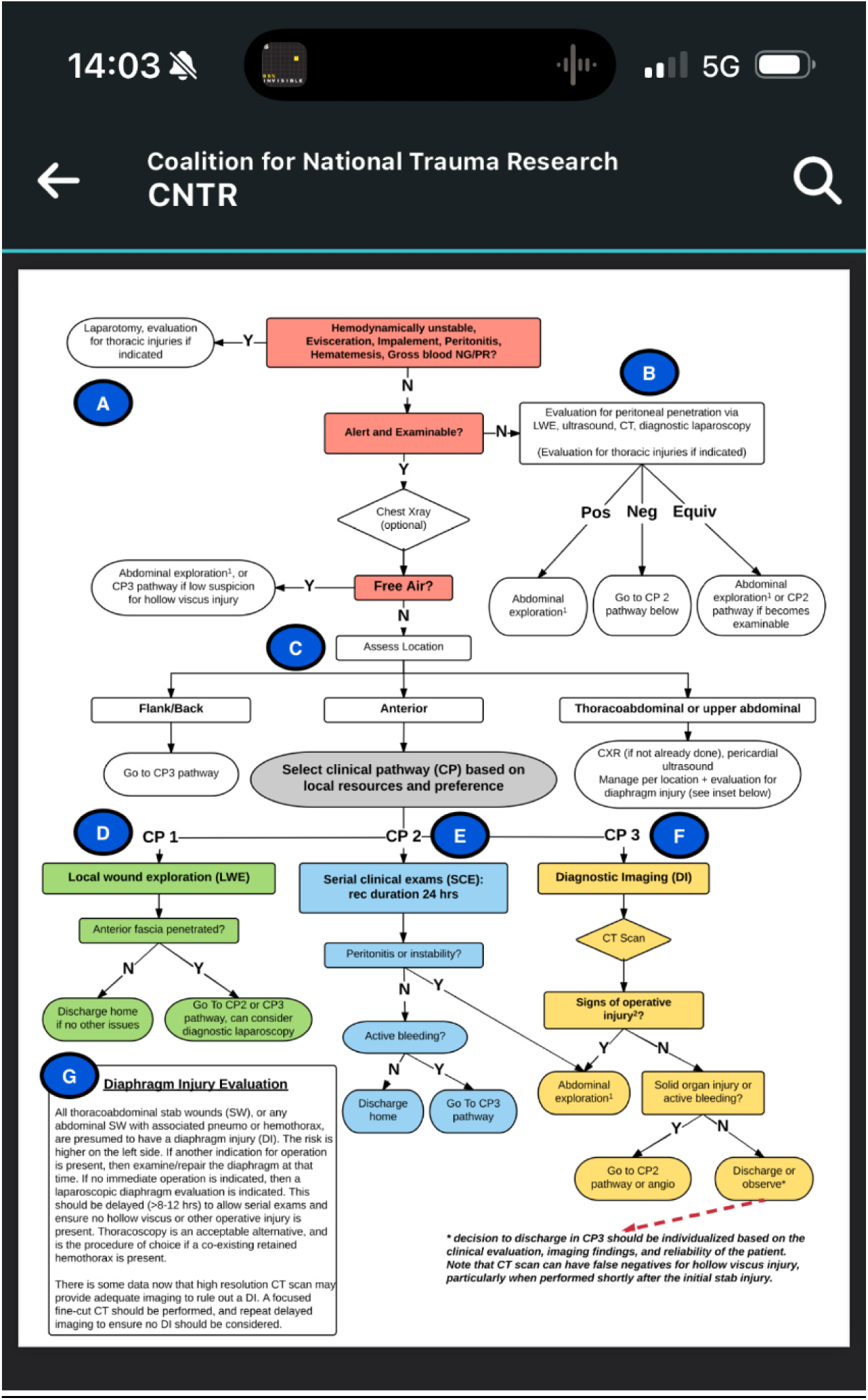

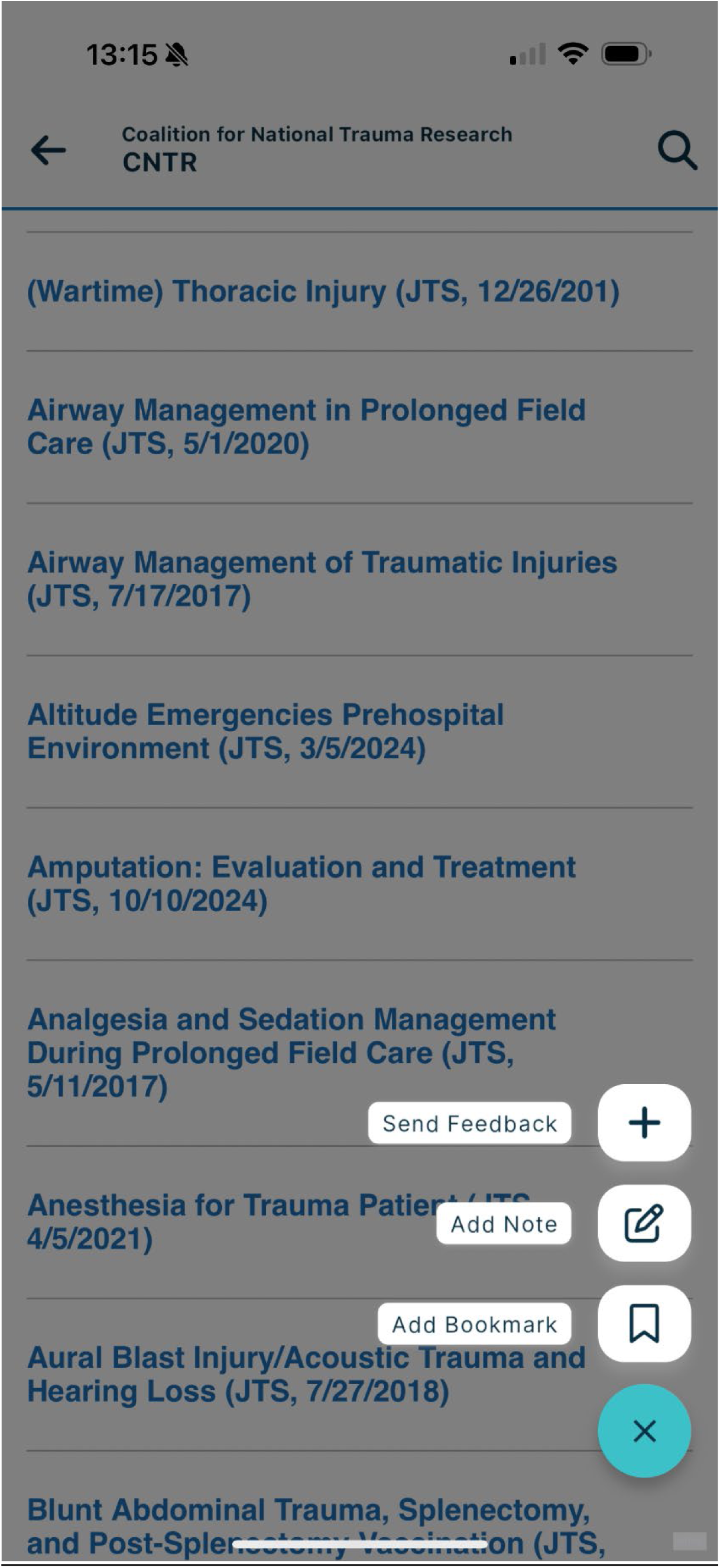

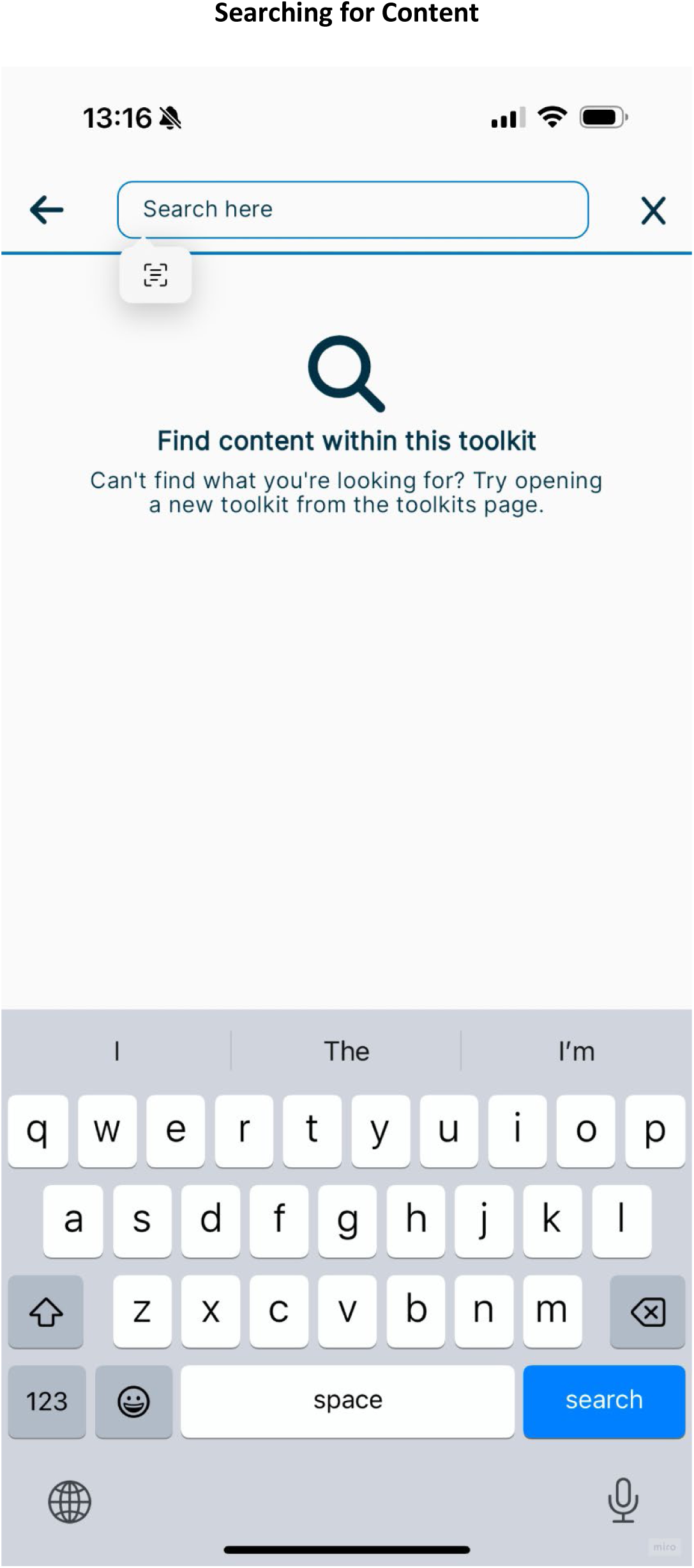

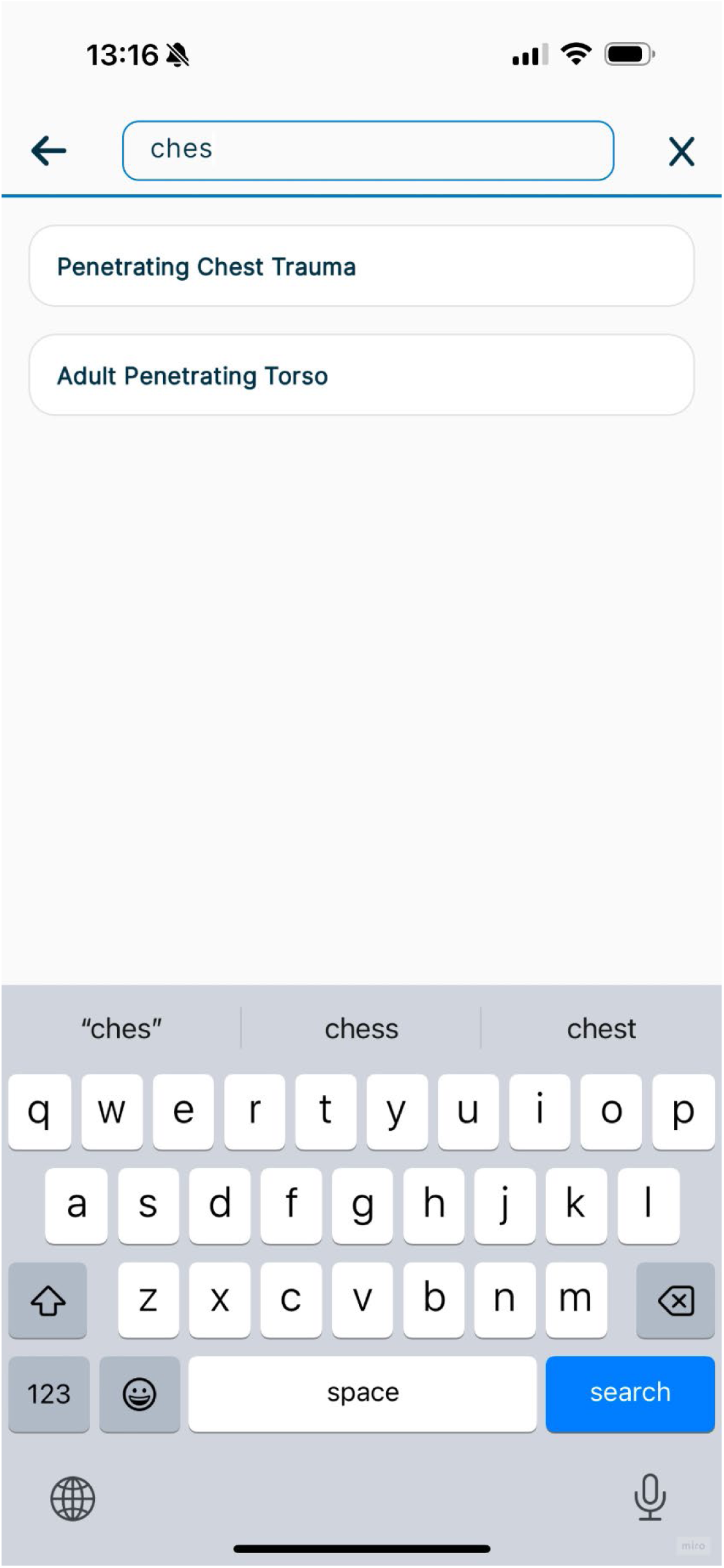

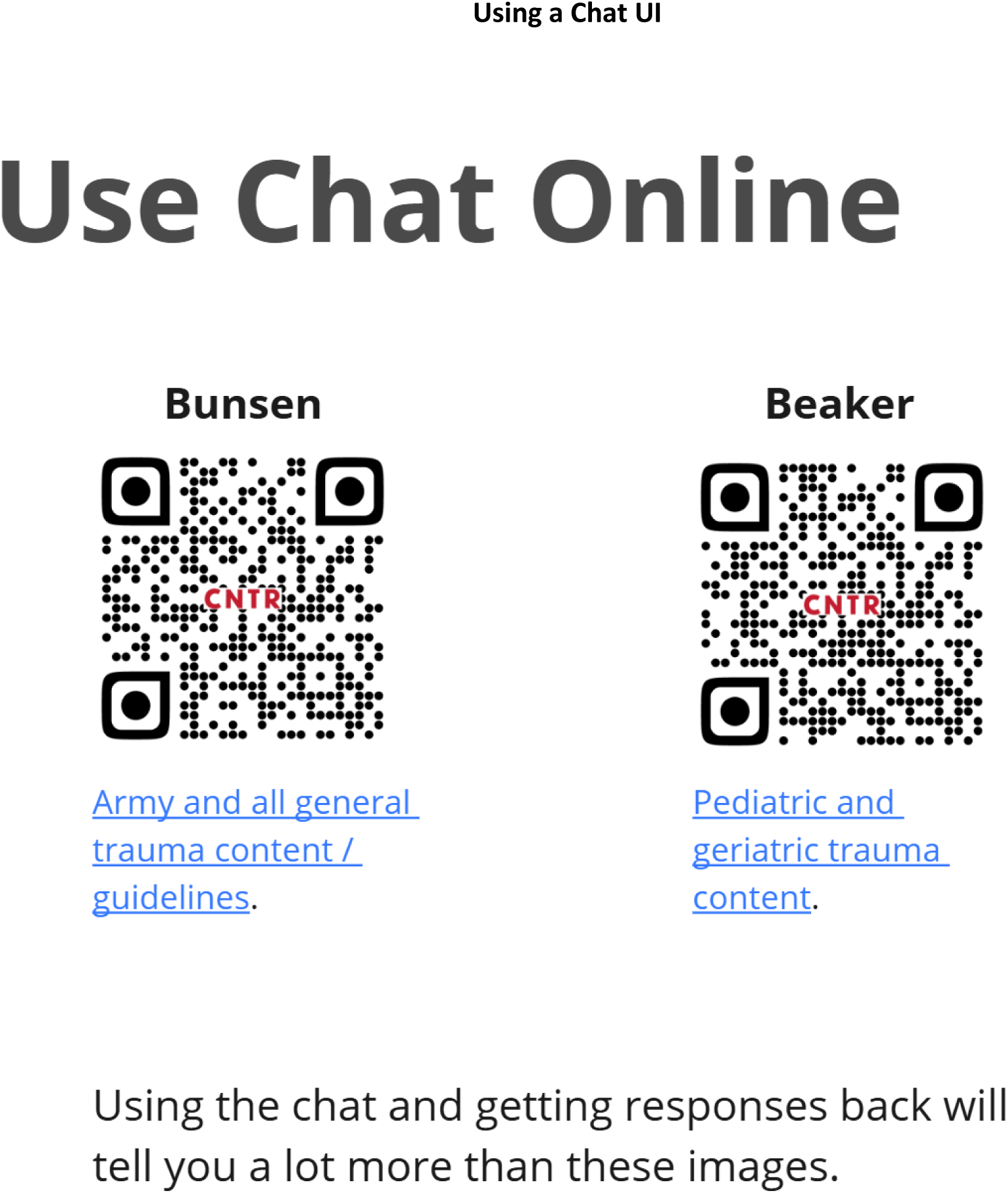

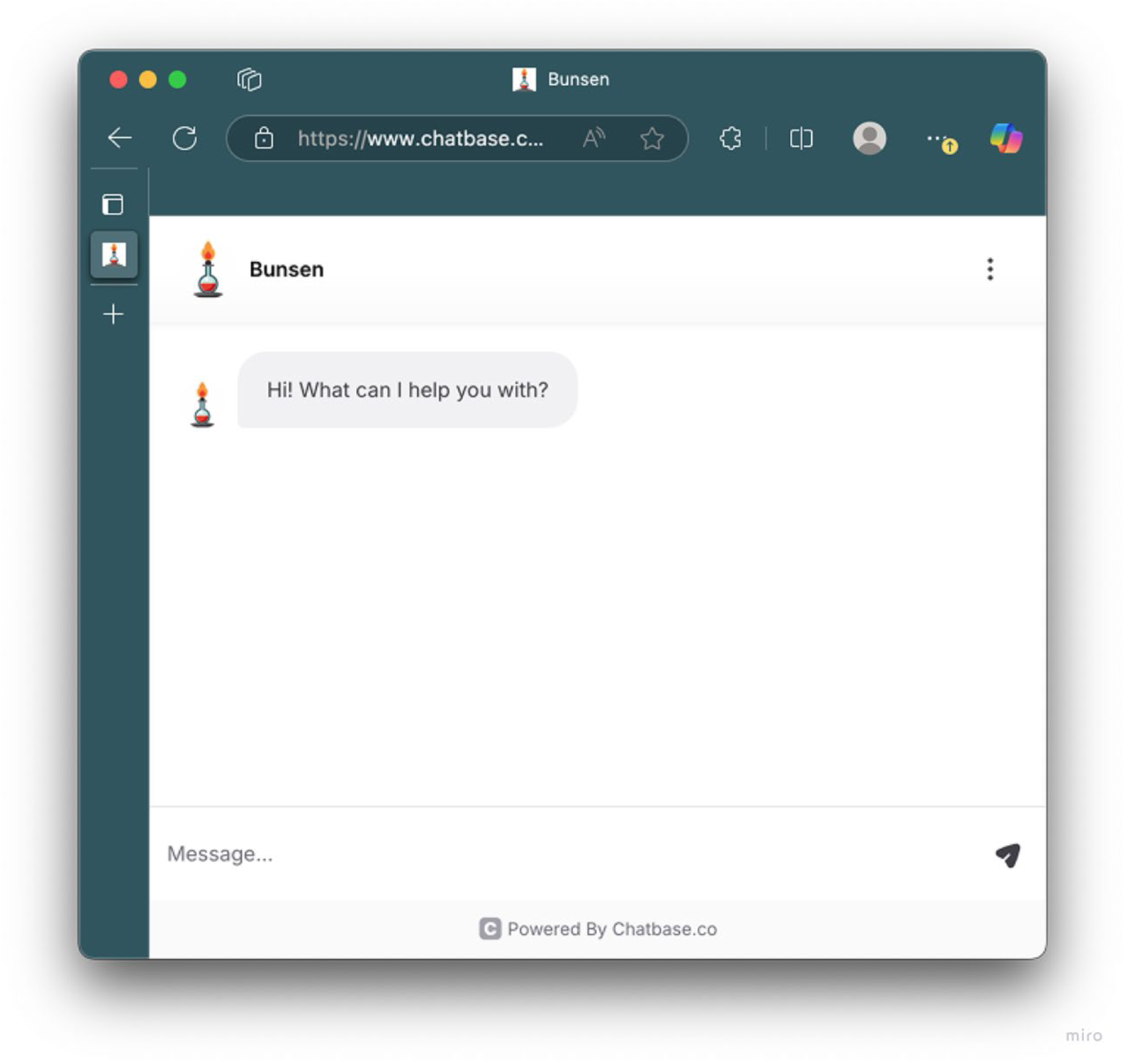

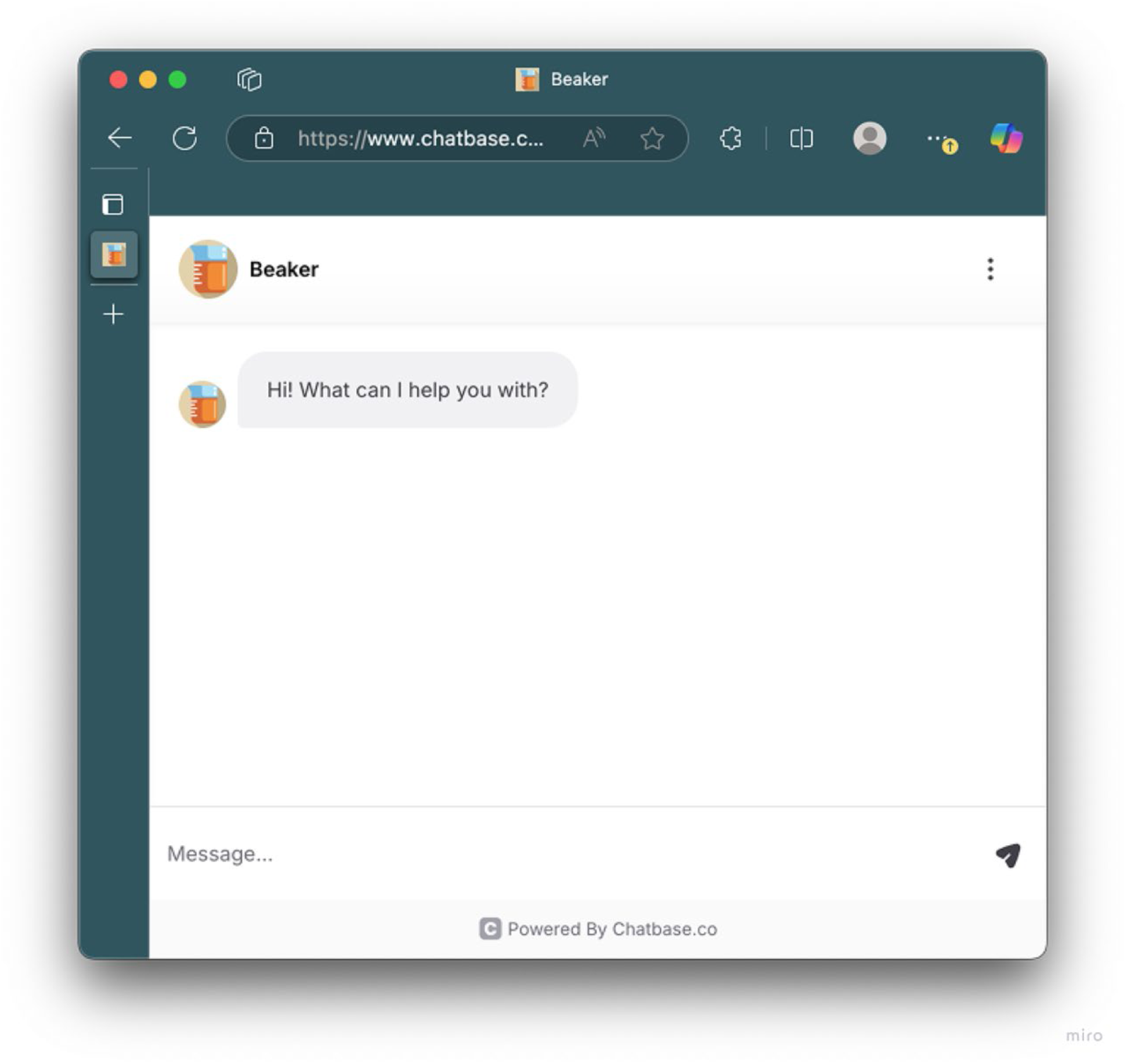

